# Unpacking Neuropsychiatry and Behavioural Neurology Training: A Scoping Review of Core Syllabus Components

**DOI:** 10.1101/2025.07.10.25329509

**Authors:** Keishema Kerr, Lauren Burns, Sheldon Benjamin, Eileen Joyce, Jasvinder Singh, Jesús Ramírez-Bermúdez, Biba Stanton, Vaughan Bell

## Abstract

**Background:** Neuropsychiatry and behavioural neurology are interdisciplinary fields that bridge psychiatry and neurology. However, there is no consensus about core curriculum components for training in these areas. This scoping review examines existing syllabuses to identify common course components and core themes in neuropsychiatry and behavioural neurology education.

**Methods:** A scoping review of both peer-reviewed and grey literature was conducted to identify unique syllabuses for neuropsychiatry and behavioural neurology courses which were analysed to determine the scope of covered disorders, conceptual approaches to training, assessment and intervention components, clinical experience requirements.

**Results:** A total of 23 unique syllabuses were identified including syllabuses from Australia, Argentina, Chile, Mexico, New Zealand, South Africa, USA, UK, and one explicitly international in scope. The analysis revealed comprehensive coverage of major neuropsychiatric conditions. Course content extended beyond the co-presentation of psychiatric and neurological disorders to include functional, behavioural, and cognitively defined disorders. Training in neuropsychiatry and behavioural neurology encompassed fundamental issues in mind and brain medicine, neuropsychology, philosophy, ethics, and social sciences. Assessment and intervention components emphasized clinical skills and knowledge relevant to both mental health and neurological services, with a particular focus on case management within social and institutional contexts.

**Conclusions:** Neuropsychiatry and behavioural neurology training integrates a broad spectrum of knowledge and skills, is aimed at a range of professionals, and is delivered as both specialist training and as embedded components within core training.

## Introduction

Neuropsychiatry and behavioural neurology are often considered sister disciplines and involve the treatment, prevention, and research of problems at the interface of psychiatry and neurology (Arciniegas & Kaufer, 2006). Psychiatric and neurological disorders frequently cooccur and co-present to clinical services. The incidence and prevalence of neurological disorders is markedly raised in patients with psychiatric disorders (Hesdorffer, 2016; Nuyen et al., 2006). Conversely, up to half of referrals to neurology clinics meet the diagnostic criteria for a neuropsychiatric diagnosis (Carson et al., 2000; Jabbar et al., 2014). Although some co-occurring difficulties may be easily managed by specialists in either discipline, consultation with, or direct management by, clinicians with specialist training in neuropsychiatry is considered best practice or, where best practice is not defined, desirable (Agrawal et al., 2008; Benjamin, 2024; Sachdev, 2005). We note here that the term ‘neuropsychiatry’ is sometimes used in a wider sense to mean the equivalent of ‘biological psychiatry’ – a field that emphasises neurobiological explanations for psychiatric disorders more widely. Here, however, we are using the term in its more common sense to mean the field that deals with conditions that lie at the intersection of psychiatry and neurology (Lishman, 1992; Ramírez-Bermúdez, Juarez, et al., 2024).

One challenge in recommending specialist training is that there is no consensus about the components of neuropsychiatry and behavioural neurology training with different authors and organisations citing different requirements for what they consider adequate training (Perez, 2024). Some authors have emphasised an approach that prioritises psychiatric complications of traditionally neurological disorders (Agrawal, 2004) with others highlighting a wider scope, including areas such as functional neurological disorder, neurodevelopmental disorders, and cognitive impairment (Arciniegas & Kaufer, 2006; Bateman et al., 2024).

Some have argued for research training as core to education in the discipline (Molina-Ruiz et al., 2024; Sachdev & Mohan, 2017) while others have focused largely on clinical competencies (Shalev & Jacoby, 2019).

There have been a variety of proposals for the place of neuropsychiatry training within medical training as a whole. Suggestions have ranged from encouraging specialists in neuropsychiatry to fully qualify as both psychiatrists and neurologists (history in Benjamin, 2024), formalising the field as a subspeciality (Silver, 2006), integrating the content as a core competency in residency training (Benjamin et al., 2014; Torous et al., 2015), to a complete ‘vertical integration’ at all stages of medical training (Mitchell & Agrawal, 2005).

Understanding shared and common components of neuropsychiatry and behavioural neurology training programs would outline a core curriculum as it is currently structured and taught. However, an important obstacle to doing this is that while some curriculum components are published within the peer-review literature, others exist solely as grey literature – either online as part of public-facing course documentation or, in some cases, solely as internal institutional documents.

One way of addressing these obstacles is to use a scoping review, which uses a systematic literature search and grey literature searches to map existing literature on the topic (Munn et al., 2018). Consequently, we used a scoping review to understand the common components of neuropsychiatry and behavioural neurology syllabuses across the world to better understand what defines neuropsychiatry as taught to trainees. We deliberately selected an inclusive approach that included academic and clinical neuropsychiatry syllabuses, the latter regardless of target profession, with the aims of clearly distinguishing these characteristics in the analysis.

## Methods

This scoping review was conducted in line with the Preferred Reporting Items for Systematic reviews and Meta-Analyses extension for Scoping Reviews (PRISMA-ScR) guidelines (Tricco et al., 2018).

### Protocol and registration

The scoping review protocol was pre-registered with INPLASY in September 2023 (registration number INPLASY202390090): https://inplasy.com/inplasy-2023-9-0090/

### Inclusions / exclusion criteria

We included any paper, study or document giving syllabus contents for a neuropsychiatry and/or behavioural neurology training course. No eligibility limits were imposed for date of the document or language. We excluded any commentaries, opinion pieces, or papers about training that did not suggest syllabus contents.

### Information sources

Three research and clinical literature databases (Pubmed, EMBASE and CINAHL) were searched on 1^st^ September 2023. The electronic database search was supplemented by reviewing reference lists. Given that many syllabus outlines may not exist in the peerreviewed literature, we also supplemented the literature database search by searching the web, and we published a request via the Global Neuropsychiatry Group to identify grey literature with syllabus contents. We requested anyone with knowledge of neuropsychiatry or behavioural neurology training courses to inform the researchers. Course leads were then contacted to request syllabus descriptions and courses were identified online to download syllabus descriptions.

### Search

For literature databases, we used the following search terms:

> “*(neuropsychiatr^*^ OR neuro-psychiatry OR neuro-psychiatric OR behavioural neurology OR behavioral neurology AND (syllabus OR curriculum OR training)”*

suitably adapted for each database. We additionally searched the web using similar search terms. A request for syllabuses to the Global Neuropsychiatry Group discussion group was included as a message to all members.

### Selection of sources of evidence

Reference from the search of literature databases were uploaded to Rayyan – an online platform which allows reviewers to collaborate and organise papers to conduct systematic style reviews. Duplicate records were identified and removed using Rayyan’s inbuilt function. Titles and abstracts were independently screened by authors KK and VB and disagreements resolved through consensus. Syllabus documents identified through the grey literature identification process was entered directly into the full-text screening stage. The full text of papers and documents was independently assessed by the same authors and the final list of documents was identified.

### Data synthesis

Syllabus components were extracted in an iterative process where individual syllabus items were extracted document by document on a coding spreadsheet. If a syllabus item was encountered that was common to a previous code, counted under this code. If it was a new item, it was added as a new code and previous documents were checked to ensure that it had not been missed previously. Initial coding was completed by author KK and checked by VB. The final data extraction sheet is included in Table S5 of the supplementary material.

Higher level syllabus component categories were coded using the upward coding approach, which involves grouping individual components into semantic categories and transforming the data from a lower to a higher level of abstraction (Jansen, 2010). Syllabus components were included into higher level categories non-exclusively, so for example the syllabus component “Stroke (and the neuropsychiatric sequelae of”) contributed to both the “Stroke” and “Secondary psychiatric syndromes” categories. We subsequently calculated frequency tables to identify how frequently syllabus component categories appeared across syllabus documents.

## Results

We identified 23 neuropsychiatry and behavioural neurology syllabus documents. The PRISMA diagram for document identification process in the study is illustrated in Figure 1. Details of all syllabus documents included in the final analysis are listed in in Table 1. We identified syllabus documents from USA (k = 8), UK (k = 7), Australia (k = 2), Australia and New Zealand (k = 1), Argentina (k = 1), Chile (k = 1), Mexico (k = 1), South Africa (k = 1) and international (k = 1).

**Table 1.** Syllabus documents included in scoping review.

| *Syllabus name* | *Year* | *Country* | *Source* | *Type* |
| --- | --- | --- | --- | --- |
| Bulletin of US Army Medical Core “Special Training in Neuropsychiatry” | 1946 | USA | PRL | Position paper |
| Benjamin (2004) “Educational issues in neuropsychiatry” | 2004 | USA | PRL | Position paper |
| Arciniegas & Kaufer (2006) “Core curriculum for training in behavioral neurology & neuropsychiatry” | 2006 | USA | PRL | Position paper |
| Lacy and Hughes (2006) “A neural systems-based neurobiology and neuropsychiatry course: integrating biology, psychodynamics, and psychology in the psychiatric curriculum” | 2006 | USA | PRL | Position paper |
| Vaishnavi et al. (2009) “Behavioral neurology and neuropsychiatry fellowship training: the Johns Hopkins model” | 2009 | USA | PRL | Position paper |
| Sachdev and Curriculum Committee of the International Neuropsychiatric Association (2010) “Core Curriculum in Neuropsychiatry of the International Neuropsychiatric Association” | 2010 | INT | Book | Position paper |
| Benjamin (2013) “Educating psychiatry residents in neuropsychiatry and neuroscience” | 2013 | USA | PRL | Position paper |
| Joint Royal Colleges of Physicians Training Board, Neuropsychiatry Section of Curriculum for Neurology Training | 2013 | UK | Web | Formal, superseded |
| Benjamin et al. (2014) “Neuropsychiatry and neuroscience milestones for general psychiatry trainees” | 2014 | USA | PRL | Position paper |
| Sachdev and Mohan (2017) “An International Curriculum for Neuropsychiatry and Behavioural Neurology” | 2017 | AUS | PRL | Position paper |
| United Council for Neurologic Subspecialties Behavioral Neurology and Neuropsychiatry Curriculum | 2020 | USA | PC | Formal, active |
| Sachdev and Mohan (2020) “Neuropsychiatry curriculum and key clinical competencies” | 2020 | AUS | Book | Position paper |
| British Psychological Society Qualification in Clinical Neuropsychology Handbook and Competency Framework | 2022 | UK | Web | Formal, active |
| Joint Royal Colleges of Physicians Training Board, Neuropsychiatry Section of Curriculum for Neurology Training | 2022 | UK | Web | Formal, active |
| Royal Australian and New Zealand College of Psychiatrists Advanced Training Neuropsychiatry Curriculum | 2023 | AUS/NZ | PC | Formal, proposed |
| College of Psychiatrists of South Africa Certificate of Neuropsychiatry Syllabus | 2023 | ZAF | PC | Formal, active |
| National Institute of Neurology and Neurosurgery, Postgraduate Course of Advanced Specialty in Neuropsychiatry | 2023 | MEX | PC | Formal, active |
| Argentine Neuropsychiatric Association Advanced Fellowship in Neuropsychiatry and Cognitive Neurology Syllabus | 2024 | ARG | Web | Formal, active |
| University of Birmingham MSc Clinical Neuropsychiatry | 2024 | UK | Web | Formal, active |
| King’s College London MSc Clinical Neuropsychiatry Syllabus | 2024 | UK | PC | Formal, active |
| University of Chile, Diploma in Adult Clinical Neuropsychiatry | 2024 | CHL | Web | Formal, active |
| Association of British Neurologists Advanced Clinical Fellowship in Behavioural Neurology and Neuropsychiatry | 2024 | UK | PC | Formal, active |
| Royal College of Psychiatrists, Syllabus for Higher Training in Neuropsychiatry | 2024 | UK | Web | Informal, active |
| ARG = Argentina; CHL = Chile; INT = International; MEX = Mexico; NZL = New Zealand; UK = United Kingdom; USA = United States of America; ZAF = South Africa; PRL = peer-review literature; PC = personal communication |  |  |  |  |

**Figure 1.**
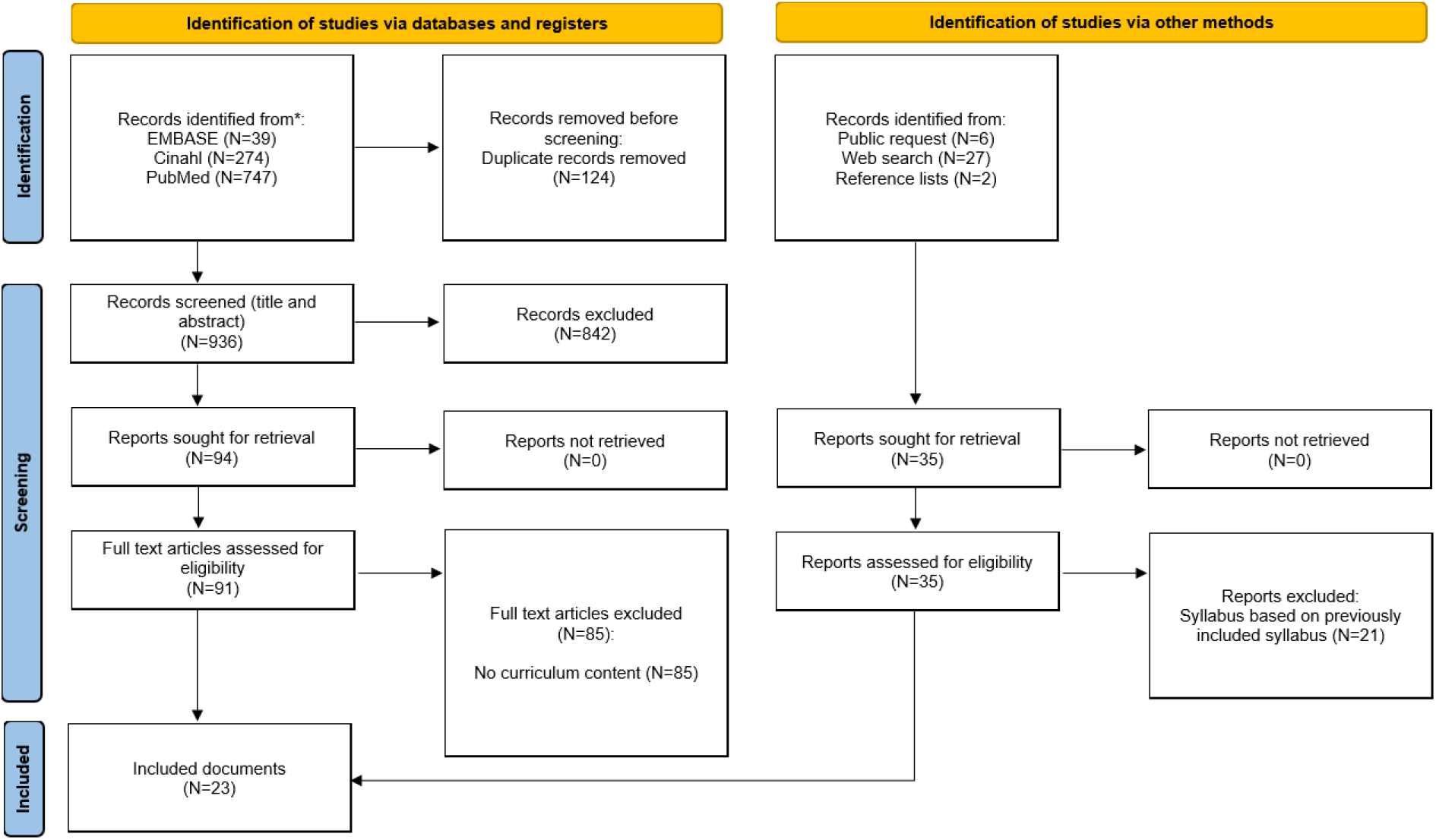
PRISMA diagram for scoping review process

### Syllabus components

Frequency tables for syllabus components and their frequency in included syllabus documents are included in Tables 2, 3, 4 and 5, highlighting topic groupings for neuropsychiatric disorders, fundamental issues, assessment and intervention. Full details of syllabus categories and their constituent syllabus components are included in Tables S1 to S4 of the supplementary material.

**Table 2.** Frequency of neuropsychiatric disorders in included syllabuses.

| *Syllabus Component* | *Syllabus count* |
| --- | --- |
| Dementia | 22 |
| Neurodevelopmental disorders | 22 |
| Movement disorders | 21 |
| Secondary psychiatric syndromes | 21 |
| Mood disorders | 21 |
| Neurobehavioural syndromes and challenging behaviour | 21 |
| Sleep disorders | 20 |
| Epilepsy and seizures | 19 |
| Cognitive impairment, selective and general | 19 |
| Acquired brain injury | 18 |
| Cerebrovascular disorders | 18 |
| Functional neurological disorders | 17 |
| Psychotic spectrum disorders | 17 |
| Demyelinating disorders | 16 |
| Neuroinfections | 16 |
| Anxiety and stress disorders | 16 |
| Substance use and addiction | 15 |
| Impulse control disorders | 14 |
| Acute and chronic pain disorders | 10 |
| Diagnosis of delirium | 10 |
| Neurooncological disorders | 9 |
| Disorders of arousal (e.g. coma, vegetative states etc) | 9 |
| Headache and migraine | 9 |
| Malingering and factitious disorders | 8 |
| Toxic exposures / ingestions | 6 |
| Hydrocephalus | 5 |
| Metabolic disorders | 5 |
| Neuroendocrine disorders | 5 |
| Sexual disorders | 4 |
| Motor neurone disease | 4 |
| Diagnosis of occupational exposure-related syndromes | 2 |
| Chronic fatigue | 2 |
| Congenital disorders | 1 |
| Nutritional disorders and nutrition in neuropsychiatry | 1 |
| Cerebrospinal fluid disorders | 1 |

**Table 3.** Frequency of fundamental issue topics in syllabuses.

| *Syllabus Component* | *Syllabus count* |
| --- | --- |
| Neuroanatomy | 21 |
| Neuropsychology and cognition | 21 |
| Neurobiology | 19 |
| Psychiatry | 17 |
| Knowledge of wider specialities | 14 |
| Research methods | 12 |
| Philosophy and ethics of neuropsychiatry | 12 |
| Professional practice issues | 12 |
| Genetics | 11 |
| Social issues and context | 10 |
| Neurology | 7 |
| Neurosurgery | 2 |
| Psychodynamics | 1 |

**Table 4.** Frequency of assessment topics in syllabuses.

| *Syllabus Component* | *Syllabus count* |
| --- | --- |
| Neuroimaging | 23 |
| Diagnosis | 23 |
| Neuropsychiatric assessment | 22 |
| Cognitive assessment | 21 |
| Neurological examination | 21 |
| EEG and electrophysiology | 19 |
| Lab tests | 15 |
| Forensic and risk assessment | 9 |
| Visual system assessment | 8 |

**Table 5.**
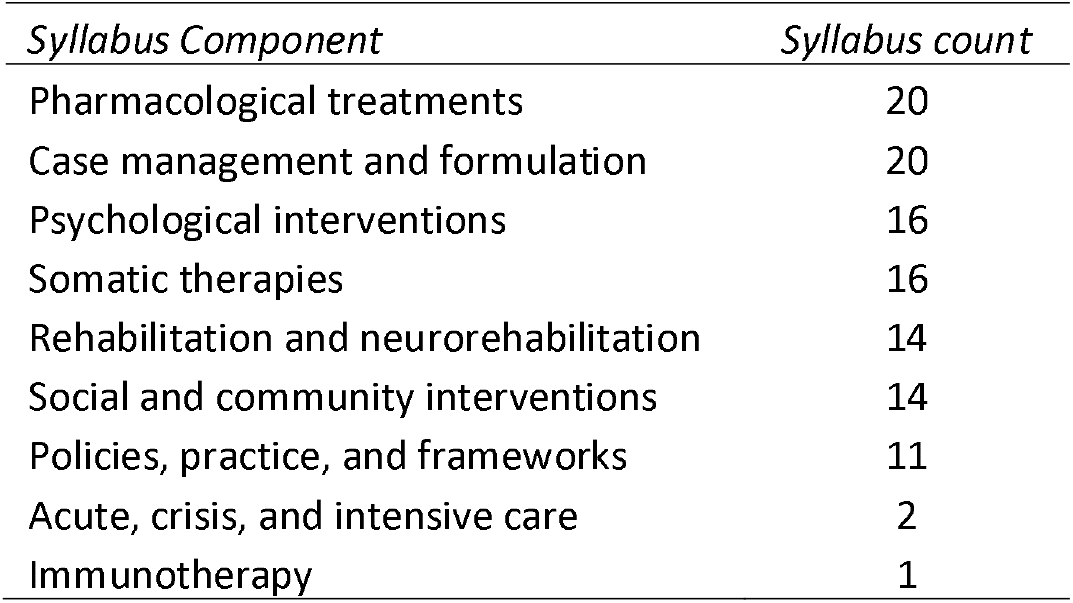
Frequency of intervention topics in syllabuses.

| *Syllabus Component* | *Syllabus count* |
| --- | --- |
| Pharmacological treatments | 20 |
| Case management and formulation | 20 |
| Psychological interventions | 16 |
| Somatic therapies | 16 |
| Rehabilitation and neurorehabilitation | 14 |
| Social and community interventions | 14 |
| Policies, practice, and frameworks | 11 |
| Acute, crisis, and intensive care | 2 |
| Immunotherapy | 1 |

### Syllabus formats

Documents included position papers from the peer-reviewed literature (k = 8), syllabuses downloaded from websites (k = 7), syllabuses received through personal communication (k = 6), and position papers published as academic books chapters (k = 2). Documents described active training courses (k = 11), a formal syllabus for an active training course that had been superseded by a subsequent syllabus (k = 1), a formal syllabus for proposed training courses that had not yet been implemented (k = 1). Of these formal training courses, k = 7 were only open to physicians, k = 1 only to clinical psychologists, k = 2 only to neurologists, k = 3 only to psychiatrists, and k = 4 were academic courses that were open to graduates including those without clinical training. Within all courses, k = 8 involved supervised clinical practice, with clinical components of these clinical training courses outlined in Table 6.

**Table 6.** Clinical training components of neuropsychiatry training course.

| *Syllabus name* | *Clinical training details* |
| --- | --- |
| Royal Australian and New Zealand College of Psychiatrists Advanced Training Neuropsychiatry Curriculum | 2-year fellowship program in a largely or exclusively neuropsychiatric service, or previous 3-year training program in a centre that offers training in both neurology and psychiatry. |
| National Institute of Neurology and Neurosurgery (Mexico), Postgraduate Course of Advanced Specialty in Neuropsychiatry | 1 year fellowship program with 3 x 4-month rotations in i) outpatient neuropsychiatry, ii) liaison, covering neurology, neurosurgery, emergency department and intensive care, and iii) inpatient psychiatry, including patients with primary psychiatric disorders, and psychiatric disorders due to neurological disease. |
| United Council for Neurologic Subspecialties Behavioral Neurology and Neuropsychiatry (USA) | 12 months supervised care of patients with focal neurobehavioural syndromes, major neuropsychiatric syndromes, and/or cognitive, emotional, and behavioural manifestations of neurological conditions. Services placement at the discretion of the program director and may include outpatient, inpatient, emergency department, other clinic types. |
| College of Psychiatrists of South Africa Sub-Specialty Certificate in Neuropsychiatry | At least 18 months fulltime experience within an accredited neuropsychiatry unit under the direction of a registered neuropsychiatrist. |
| Royal College of Psychiatrists (UK) | No formal training pathway but 1 year training posts offered as part of integrated residency training or standalone training post. |
| Federation of the Royal Colleges of Physicians of the UK Joint Royal Colleges of Physicians Training Board, Neurology Training Curriculum (2013, 2022) | No specific neuropsychiatry placement required but requires neuropsychiatry competencies to be completed as part of neurology residency. |
| Association of British Neurologists Advanced Clinical Fellowship in Behavioural Neurology and Neuropsychiatry (Proposed) | 12 months supervised clinical practice, involving brain injury, inpatient neuropsychiatry, general liaison neuropsychiatry, and old age liaison neuropsychiatry. In addition, attendance at weekly multidisciplinary team meetings, including neuropsychiatry-focused radiology and EEG, brain injury, and less frequently complex somatic symptoms and post-COVID syndrome. |
| British Psychological Society Qualification in Clinical Neuropsychology | 24 months clinical practice in neuropsychology achieving a range of neuropsychology competencies that include neuropsychiatry competencies. Not service specific. |

## Discussion

In this scoping review of both peer-reviewed and grey literature, we identified 23 unique syllabuses for neuropsychiatry and behavioural neurology courses to identify common course components. The identified course components indicate a thorough coverage of major neuropsychiatric conditions with the most prevalent disorders covered by most courses and lower prevalence disorders covered less commonly. The covered disorders do not solely focus on the co-presentation of traditionally psychiatric and neurological disorders, but also include functional, behavioural and cognitively defined disorders. Neuropsychiatry and behavioural neurology training was conceptualised as teaching across a range of fundamental issues across mind and brain medicine, neuropsychology, differing degrees of philosophy, ethics and social science. Assessment and intervention components focused on a range of clinical skills and knowledge necessary to work across both mental health and neurological services, with a focus on management of cases in the social and institutional context of the trainee.

We note a clear evolution from proposals in the earlier peer-reviewed literature concerning the necessary components of training to the later development of formal training courses – likely indicating a healthy progression from debate to implementation. Of the syllabuses from formal training programmes, we also note that neuropsychiatry and behavioural neurology training is quite diverse in terms of how it is positioned with regard to clinical training pathways and academic learning. Some training is profession specific – exclusively aimed at physicians, specific medical specialties or clinical psychologists – some is embedded as a component within wider core clinical training, some implements training for a clinical specialisation, while some is designed as academic study, aimed at developing a domain knowledge and is inclusive of wider health professionals and even those with a purely academic interest.

We also note what may at first seem like an anomaly in the frequency of syllabus components. In the ‘fundamental issues’ section the specifically labelled ‘psychiatry’ component appears more frequently than the ‘neurology’ component. However, this appears to be down to naming conventions rather than content per se. There were more syllabuses that classified themselves as ‘neuropsychiatry’ rather than ‘neuropsychiatry and behavioural neurology’ or solely ‘behavioural neurology’, and so some topics were preferentially labelled as ‘psychiatry of’ or ‘neuropsychiatry of’ whereas core neurology components tended not to be named ‘neurology of’. In fact, the analysis of syllabus components as a whole shows that content from psychiatry, neurology, neuropsychology, and social science is widely represented. It is also worth noting a broader reality, that in the majority of countries, formal training routes in neuropsychiatry and behavioural neurology are not available although trainees report varying levels of integration into core training but express a clear demand for further exposure (Costello et al., 2025; Molina-Ruiz et al., 2024).

Combining internationally sourced syllabus components risks implying that neuropsychiatry and behavioural neurology should be defined as a ‘global average’. While some common components are likely to be universal, the profile reported should not be seen as prescriptive. The extent to which neuropsychiatry and behavioural neurology needs to be adapted to local or regional contexts has been debated with regard to the needs of Latin America (e.g. Ramírez-Bermúdez et al., 2024), and India and South Asia (Krishnamoorthy & Misra, 2020).

For example, the neuropsychiatry of neglected tropical diseases (Berkowitz et al., 2015) is a common focus of clinical and research concern and high priority in some regions (Rosa et al., 2025) but less so in others. Therefore, any standardised framework should remain flexible, allowing for core components that transfer across countries while including cultural, clinical, and educational adaptations that reflect the priorities of different regions, and the needs of different professions within those regions.

We highlight some potential limitations of this scoping review. Although we did not limit syllabuses by language, database searches were completed in English. The databases we selected have a range of non-English language journals abstracted in English, although it is likely that non-English language journals are less likely to indexed in these databases at all. We complemented our database searches by including a request to the Global Neuropsychiatry Group discussion network which, at the time of the query, included about 300 people from across the world with self-described interest in neuropsychiatry, although because this is primarily an English language group and relies on motivated individuals to respond, a selection bias may have occurred. We identified only three non-English language syllabuses, all from Latin America, but it is unclear whether this reflects a greater presence of neuropsychiatry and behavioural neurology training in the region or simply greater visibility to our search methods. We also note that there is no standard format for the communication of syllabuses and the documents retrieved varied in their format – from formal descriptions designed to satisfy the accreditation criteria of academic or regulatory bodies to online descriptions aimed at prospective trainees. This may have under-recognised smaller topics, as these were less likely to be included in documents aimed at the public, which appeared more focused on ‘headline’ topics.

In conclusion, we report a clear development of neuropsychiatry and behavioural neurology training from early proposals to formal syllabuses. These courses integrate a broad spectrum of knowledge from psychiatry, neurology and neuropsychology, and are delivered in a number of diverse formats, variously aimed at specific professions or as broad academic study.

## Data Availability

This is a scoping review and so all data is referenced in the manuscript

## Ethics statement

As a review of published literature, this scoping review was exempt from ethical review.

## Author Contribution Statement

KK, LL and VB were involved in developing the research question, designing the study, and carrying it out. All authors were involved in interpreting the data and writing the article.

## Transparency Declaration

The lead author and manuscript guarantor (VB) affirms that the manuscript is an honest, accurate, and transparent account of the study being reported; that no important aspects of the study have been omitted; and that any discrepancies from the study as planned (and, if relevant, registered) have been explained.

