## Supplementary material for "Unpacking Neuropsychiatry and Behavioural Neurology Training: A Scoping Review of Core Syllabus Components"

**What are the components of neuropsychiatry and behavioural neurology training courses? A scoping review**

**Table S1.** Syllabus items for the neuropsychiatric disorders categories

| *Syllabus component* | *Syllabus item* |
| --- | --- |
| Dementia | - Diagnosis and management of neuropsychiatric aspects of movement disorders (including idiopathic Parkinson’s disease, progressive supranuclear palsy, corticobasal syndromes, dementia with Lewy Bodies, Huntington’s disease, dystonia, Tourette’s syndrome) - Dementia - Neurodegenerative disorders - Alzheimer's disease - Frontotemporal Dementia - Lewy body disease - Parkinson's disease - Huntington's disease - Biomarkers of neurobehavioral and neuropsychiatric conditions, including neurodegeneration and neurotrauma - Neurocognitive disorders, including delirium, mild cognitive impairment (i.e., Mild Neurocognitive Disorder) and dementia (i.e., Major Neurocognitive Disorder) - Chronic / progressive behavioural and cognitive change |
| Acquired brain injury | - Brain injury - Diagnosis and management of neuropsychiatric aspects of brain injury - Assessment of patients with acquired brain injury in acute inpatient and specialist outpatient settings, including neurological, cognitive and behavioural aspects of the presentation - Ability to support other professionals with challenging aspects of the neuropsychiatry of brain injury, including managing challenging behaviour and assessing mental capacity - Acquired brain injury (both traumatic and non-traumatic) - Traumatic brain injury (and sequelae) - Hypoxic-ischemic brain injury - Biomarkers of neurobehavioral and neuropsychiatric conditions, including neurodegeneration and neurotrauma - Understanding of the range of cognitive and behavioural deficits associated with acquired brain injury, and the impact of these on quality of life and care-giver burden |
| Cerebrovascular disorders | - Stroke (and its neuropsychiatric sequelae) - Cerebrovascular disorders - Hypoxic-ischemic brain injury - Vascular disorders |
| Epilepsy and seizures | - Epilepsy - Assessment of patients with seizures complicated by cognitive or psychiatric symptoms and identify the contributions of seizure activity, medication and co-morbid mental health conditions to their difficulties - Management of the ongoing care of patients with epilepsy complicated by psychiatric comorbidity or additional functional / non-epileptic seizures - Seizure disorders - Medico-legal aspects of epilepsy - Diagnosis and management of neuropsychiatric aspects of epilepsy - Understanding of the range of cognitive symptoms and psychiatric co-morbidity associated with epilepsy (including mood disorders and psychosis) and anti-epileptic drugs |
| Functional neurological disorders | Pathophysiology of functional symptoms   - Use of clinical assessment to make an accurate diagnosis of patients referred to a functional disorders clinic, including assessment of co-existing functional and “organic” symptoms, psychiatric co-morbidity and multi-system functional somatic symptoms - Functional Neurological Disorder (FND) (also cited as conversion disorder) - Use of clinical skills to differentiate functional versus “organic” movement disorder, including in challenging cases where both may co-exist - Management of the ongoing care of patients with epilepsy complicated by psychiatric comorbidity or additional functional / non-epileptic seizures - Complex somatic and medically unexplained symptoms - Dissociative disorder |
| Movement disorders | - Diagnosis and management of neuropsychiatric aspects of movement disorders (including idiopathic Parkinson’s disease, progressive supranuclear palsy, corticobasal syndromes, dementia with Lewy Bodies, Huntington’s disease, dystonia, Tourette’s syndrome) - Use of clinical skills to differentiate functional versus “organic” movement disorder, including in challenging cases where both may co-exist - Use of clinical assessment to diagnose common psychiatric complications of movement disorders including anxiety, depression, impulse control disorders and psychosis - Parkinson's disease - Huntington's disease - Motor disorders (including developmental coordination disorder and stereotypic movement disorder) - Catatonic behaviour and other motoric disturbances - Startle syndromes and myoclonus - Tic disorders (including Gilles de la Tourette's) |
| Secondary psychiatric syndromes | - Diagnosis and management of neuropsychiatric aspects of movement disorders (including idiopathic Parkinson’s disease, progressive supranuclear palsy, corticobasal syndromes, dementia with Lewy Bodies, Huntington’s disease, dystonia, Tourette’s syndrome) - Diagnosis and management of neuropsychiatric aspects of brain injury - Understanding the range of cognitive and behavioural deficits associated with acquired brain injury, and the impact of these on quality of life and care-giver burden - Assessment of patients with acquired brain injury in acute inpatient and specialist outpatient settings, including neurological, cognitive and behavioural aspects of the presentation - Diagnosis and management of neuropsychiatric aspects of epilepsy - Understanding of the range of cognitive symptoms and psychiatric co-morbidity associated with epilepsy (including mood disorders and psychosis) and anti-epileptic drugs - Assessment of patients with seizures complicated by cognitive or psychiatric symptoms and identify the contributions of seizure activity, medication and co-morbid mental health conditions to their difficulties - Management of the ongoing care of patients with epilepsy complicated by psychiatric comorbidity or additional functional / non-epileptic seizures - Use of clinical assessment to diagnose common psychiatric complications of movement disorders including anxiety, depression, impulse control disorders and psychosis - Psychotic symptoms in patients with neurological disorders - Secondary psychiatric disorders, i.e. psychosis, depression, mania and anxiety disorders secondary to ‘organic’ brain disease - Substance-induced psychiatric disorders – alcohol, drugs of abuse, addiction etc - Focal neurobehavioural syndromes - Stroke (and the neuropsychiatric sequelae of) - Neuropsychiatry of substance abuse |
| Demyelinating disorders | - Multiple sclerosis (and other demyelinating diseases) - Neuromuscular disease - Infections and related inflammatory disorders |
| Neuroinfections | - Autoimmune disorders affecting the central nervous system, including autoimmune encephalopathies - Post-COVID syndrome - CNS infections (including neuropsychiatric manifestations of HIV) - Infections and related inflammatory disorders |
| Neurodevelopmental disorders | - Autism - Attentional disorders (adult ADHD and related syndromes) - Tic disorders (including Gilles de la Tourette's) - Developmental disorders - Intellectual disabilities - (Specific) learning disorders - Behavioural disorders (in intellectual disability, autism, and other developmental disabilities) - Motor disorders (including developmental coordination disorder and stereotypic movement disorder) |
| Cognitive impairment, selective and general | - Cognitive disorders - Understanding of the range of cognitive and behavioural deficits associated with acquired brain injury, and the impact of these on quality of life and care-giver burden - Non-dementing cognitive disorders - Assessment of deficits in language, praxis, gnosis, visuospatial function and other cognitive syndromes - Frontal / executive syndromes (of disinhibitory and non-spontaneous types) - Cognitive impairment - Focal neurobehavioural syndromes - Disorders of Perception (e.g., illusions, hallucinations, sensory impairments) - Attention (e.g., delirium / acute confusional states, hemispatial inattention / neglect) - Disorders of Language (e.g., aphasias, affective aprosodias) - (Disorders of) Memory (e.g., amnesias) - Disorders of Praxis (e.g., apraxias) - Disorders of Recognition (e.g., agnosias) - (Disorders of) Executive function - Amnestic disorders - Communication disorders - (Specific) learning disorders - Disruptive, impulse-control and conduct disorders - Neurocognitive disorders, including delirium, mild cognitive impairment (i.e., Mild Neurocognitive Disorder) and dementia (i.e., Major Neurocognitive Disorder) - Disorders of Speech and Thought - Disorders of Judgment |
| Psychotic spectrum disorders | - Understanding of the range of cognitive symptoms and psychiatric co-morbidity associated with epilepsy (including mood disorders and psychosis) and anti-epileptic drugs - Use of clinical assessment to diagnose common psychiatric complications of movement disorders including anxiety, depression, impulse control disorders and psychosis - Secondary psychiatric disorders, i.e. psychosis, depression, mania and anxiety disorders secondary to ‘organic’ brain disease - Disorders of Perception (e.g., illusions, hallucinations, sensory impairments) - Psychotic symptoms in patients with neurological disorders - Neurobiology of Schizophrenia |
| Mood disorders | - Understanding of the range of cognitive symptoms and psychiatric co-morbidity associated with epilepsy (including mood disorders and psychosis) and anti-epileptic drugs - Use of clinical assessment to diagnose common psychiatric complications of movement disorders including anxiety, depression, impulse control disorders and psychosis - Psychiatric disorders including depression, anxiety disorders, PTSD, emotionally unstable personality disorder - Secondary psychiatric disorders, i.e. psychosis, depression, mania and anxiety disorders secondary to ‘organic’ brain disease - Disorders of mood - Disorders of affect |
| Malingering / factitious disorders | - Factitious disorders - Malingering |
| Substance use / addiction | - Substance-induced psychiatric disorders – alcohol, drugs of abuse, addiction etc - Neuropsychiatry of substance abuse |
| Neurobehavioural syndromes and challenging behaviour | - Aggression (and violence) - Ability to support other professionals with challenging aspects of the neuropsychiatry of brain injury, including managing challenging behaviour and assessing mental capacity - Use of clinical assessment to diagnose common psychiatric complications of movement disorders including anxiety, depression, impulse control disorders and psychosis - Acute behavioural disturbance - Chronic / progressive behavioural and cognitive change - Frontal / executive syndromes (of disinhibitory and non-spontaneous types) - Focal Neurobehavioural syndromes - Disruptive, impulse-control and conduct disorders - Paraphilic disorders - Personality disorders / personality change due to neurological conditions - Behavioural disorders (in intellectual disability, autism, and other developmental disabilities) - Disturbances of appetite and sexual behaviour - Disorders of Judgment |
| Neurooncological disorders | - Neuro-oncology - Primary and secondary brain tumours - Neuropsychology of neoplastic and systemic disorders |
| Anxiety / stress disorders | - Anxiety Disorders - Use of clinical assessment to diagnose common psychiatric complications of movement disorders including anxiety, depression, impulse control disorders and psychosis - Psychiatric disorders including depression, anxiety disorders, PTSD, emotionally unstable personality disorder - Secondary psychiatric disorders, i.e. psychosis, depression, mania and anxiety disorders secondary to ‘organic’ brain disease - Trauma - Trauma and stress-related disorders - Obsessive-compulsive and related disorders - Dissociative disorder - Depersonalisation / DID |
| Sexual disorders | - Sexual disorders - Disturbances of appetite and sexual behaviour - Paraphilic disorders |
| Impulse control disorders | - Use of clinical assessment to diagnose common psychiatric complications of movement disorders including anxiety, depression, impulse control disorders and psychosis - Disruptive, impulse-control and conduct disorders - Frontal / executive syndromes (of disinhibitory and non-spontaneous types) |
| Single item syllabus components   - Sleep disorders - Disorders of arousal (e.g., coma, vegetative states, minimally conscious states) - Hydrocephalus - Toxic exposures / ingestions - Metabolic disorders - Headache / migraine - Acute and chronic pain (disorders) - (Neuro) endocrine disorders - Diagnosis of delirium - Diagnosis of occupational exposure-related syndromes - Motor neurone disease - Congenital disorders - Nutritional disorders and Nutrition in Neuropsychiatry - Chronic fatigue - CSF disorders |  |

**Table S2.** Syllabus items for the fundamental issues categories

| *Syllabus component* | *Syllabus item* |
| --- | --- |
| Neuroanatomy | - Knowledge of the neuroscientific principles underlying neuropsychiatric practice (in relation to neuroanatomy, neurophysiology, neurochemistry and neuropharmacology) - Knowledge of brain structure at the macroscopic and microscopic levels, in particular the knowledge of neuronal networks, the limbic system, the neuroanatomical substrates of memory and the frontal executive system - Knowledge of CNS structure-function correlates - Structural and functional neuroanatomy (networks of psychiatric significance) - Cerebral cortex and its parts - Limbic and paralimbic structures - Basal ganglia - Brainstem (Hypothalamus, Thalamus and Internal Capsule) - Cerebellum - White matter - Cortico-cortical-subcortical circuits and networks - Regional cerebral specialization - Amygdala |
| Neuropsychology and cognition | - Knowledge of brain structure at the macroscopic and microscopic levels, in particular the knowledge of neuronal networks, the limbic system, the neuroanatomical substrates of memory and the frontal executive system - Knowledge of CNS structure-function correlates - Structural and functional neuroanatomy (networks of psychiatric significance) - Cognition (e.g., arousal, perception, attention, language, memory, praxis recognition, visuospatial function, executive function) - Emotion (e.g., mood, affect, prosody [affective communication]) - Behavior (e.g., motivation, comportment, personality) - Perception (e.g., illusions, hallucinations, sensory impairments) - Attention (e.g., delirium / acute confusional states, hemispatial inattention / neglect) - Social cognition (e.g., comportment, emotional recognition, theory of mind) - Motivation - Regional cerebral specialization - Neuropsychology and cognitive neuroscience - Psychobiology (biological psychology) |
| Neurobiology | - Pathophysiology of functional symptoms - Neuroscientific principles underlying neuropsychiatric practice (in relation to neuroanatomy, neurophysiology, neurochemistry and neuropharmacology) - Knowledge of clinical neuroscience - Knowledge of neurochemistry (especially neurotransmitter and receptor function, neurotransmitters, neuropeptides, neurohormones) - The biochemical basis of neuropsychopharmacology - The basic principles of neurophysiology - Neurogenetics - Clinical or research neurophysiology (including electrodiagnostic and neuromagnetic assessment techniques) - Neuropharmacology - Elemental neurological function - Neurobiology - Hypnosis - Mindfulness |
| Knowledge of wider specialities | - Old age liaison - Neuro-oncology - Support to community teams or allied disciplines treating patients - Relationship between neuropsychiatry and allied psychiatric subspecialties such as old age, child and learning disability psychiatry - Geriatric Behavioural Neurology and Neuropsychiatry - Pediatric BN & NP - Neurogenetics - Clinical or research neurophysiology (including electrodiagnostic and neuromagnetic assessment techniques) - Neuropathology - Epidemiology, public health, public policy, and / or public advocacy - Administration / administrative psychiatry - Education - Principles of neurology - Paediatrics neurology / neuroscience - Principles underlying the choice and integration of services and interventions in neuropsychiatry - Prevention and health promotion in Neuropsychiatry - Neurosurgery |
| Psychiatry | - Inpatient Neuropsychiatry - General liaison neuropsychiatry - Developmental Neuropsychiatry - Forensic neuropsychiatry - Understanding patterns of psychiatric symptomatology and presentation - Relationship between neuropsychiatry and allied psychiatric subspecialties such as old age, child and learning disability psychiatry - Geriatric Behavioural Neurology and Neuropsychiatry - Pediatric BN & NP - Administration / administrative psychiatry - Medico-legal aspects to the practice of neuropsychiatry (including the Mental Health Act) - History of neuropsychiatry - Psychopathology - Psychosomatic psychiatry - Pathophysiology of neuropsychiatric disorders - Epidemiology of neuropsychiatric disorders in various populations - Nutritional disorders and Nutrition in Neuropsychiatry - Principles underlying the choice and integration of services and interventions in neuropsychiatry - Prevention and health promotion in Neuropsychiatry - Psychiatric phenomenology - Conceptual, philosophical and legal issues in neuropsychiatry - Sociology of neuropsychiatry |
| Neurology | - Geriatric Behavioural Neurology and Neuropsychiatry - Principles of neurology - Elemental neurological function - Paediatric neurology / neuroscience - Phenomenology of neurologic disorders - Clinical or research neurophysiology (including electrodiagnostic and neuromagnetic assessment techniques) - Neuropharmacology - Neuropathology |
| Research methods | - Understanding the current guidelines and evidence base for treatments - All the steps in an empirical project - Clinical or research neurophysiology (including electrodiagnostic and neuromagnetic assessment techniques) - Critique and synthesis of the existing scientific literature and application to practice - Statistics and research methods (including critical thinking in research and scholarship) - Animal Models of Neuropsychiatric diseases |
| Philosophy and ethics of neuropsychiatry | - Ethical codes of conduct - Basic grasp of issues related to the mind-brain debate, the biology of consciousness and other neurophilosophical issues - Medico-legal aspects of epilepsy - Medico-legal aspects to the practice of neuropsychiatry (including MHA) - Principles underlying the choice and integration of services and interventions in neuropsychiatry - Conceptual, philosophical and legal issues in neuropsychiatry - Knowledge and appropriate application of adult safeguarding processes |
| Genetics | - The basic principles of genetics and immunology as they apply to the CNS - Neurogenetics |
| Social issues and context | - Epidemiology, public health, public policy, and / or public advocacy - Education - History of neuropsychiatry - Prevention and health promotion in Neuropsychiatry - Sociology of neuropsychiatry |
| Professional practice issues | - Multidisciplinary team working - Support to community teams or allied disciplines treating patients |
| Single item syllabus components   - Psychodynamics |  |

**Table S3.** Syllabus items for the assessment categories

| *Syllabus component* | *Syllabus item* |
| --- | --- |
| Cognitive assessment | - Assessment of patients with acquired brain injury in acute inpatient and specialist outpatient settings, including neurological, cognitive and behavioural aspects of the presentation - Performance of a cognitive examination (simple and extended) - Assessment of deficits in language, praxis, gnosis, visuospatial function and other cognitive syndromes - Interpretation of neuropsychological tests - Neuropsychological (psychometric) testing - General assessment of cognition, emotion, and behavior - Standardized assessments of neuropsychiatric symptoms and syndromes - Content, sensitivity, and specificity of neuropsychological testing - Factors that influence neuropsychological test performance - Relationship between neuropsychological tests and bedside or office-based quantified clinical assessments - Cognitive tests |
| Neuropsychiatric assessment | - Use of clinical assessment to make an accurate diagnosis of patients referred to a functional disorders clinic, including assessment of co-existing functional and “organic” symptoms, psychiatric co-morbidity and multi-system functional somatic symptoms - Use of clinical skills to differentiate functional versus “organic” movement disorder, including in challenging cases where both may co-exist - Use of clinical assessment to diagnose common psychiatric complications of movement disorders including anxiety, depression, impulse control disorders and psychosis - Ability to gather a neuropsychiatric history; this includes all of the information routinely gathered as part of a psychiatric and medical history, screening tests and informant questionnaires - Interpret any abnormal signs elicited in neurological / medical examination and place them in context of presentation and a differential diagnosis - Neuropsychiatric assessment - Mental Status Examination (and interpretation of) - Assessment and management of treatment of patients with neuropsychiatric disorders such as those with psychiatric and behavioural symptoms and co-existing neurological disorder - Use of a formulation-based approach to help patients (including age, gender, culture, SES, familial factors) - (Assessment of) Impact upon relatives and carers - Assessment of patients with seizures complicated by cognitive or psychiatric symptoms and identification of the contributions of seizure activity, medication and co-morbid mental health conditions to their difficulties - Ability to support other professionals with challenging aspects of the neuropsychiatry of brain injury, including managing challenging behaviour and assessing mental capacity - Functional assessments |
| Neuroimaging | - (Neuro) Radiology - Understanding of the indications for, and interpretations of, the various forms of brain imaging, both structural and functional, including MRI, CT, SPECT and PET - Neuroimaging - Principles and applications of structural and functional imaging of the brain - Correlation between neuroimaging and clinical examination - Ventriculography |
| Neurological examination | - Able to assess patients with acquired brain injury in acute inpatient and specialist outpatient settings, including neurological, cognitive and behavioural aspects of the presentation - Neurological examination - Interpretation of abnormal signs elicited in neurological / medical examination and placement of them in context of presentation and a differential diagnosis - Neurological “soft-signs” - The use of neurological examination rating scales and the interpretation of such data |
| EEG and electrophysiology | - EEG (evoked potentials and brain-mapping) and other neurophysiological investigations and their interpretation - Electrophysiologic Testing - Clinical or research neurophysiology (including electrodiagnostic and neuromagnetic assessment techniques) - Encephalograms |
| Diagnosis | - Diagnosis and management of neuropsychiatric aspects of movement disorders (including idiopathic Parkinson’s disease, progressive supranuclear palsy, corticobasal syndromes, dementia with Lewy Bodies, Huntington’s disease, dystonia, Tourette’s syndrome - Use of clinical assessment to make an accurate diagnosis of patients referred to a functional disorders clinic, including assessment of co-existing functional and “organic” symptoms, psychiatric co-morbidity and multi-system functional somatic symptoms - Diagnosis and management of neuropsychiatric aspects of brain injury - Diagnosis and management of neuropsychiatric aspects of epilepsy - Use of clinical assessment to diagnose common psychiatric complications of movement disorders including anxiety, depression, impulse control disorders and psychosis - Neuropsychiatric diagnosis including history and examination, neurophysiological investigations, neuroimaging, neuropsychology, and other investigations - Interpretation of abnormal signs elicited in neurological / medical examination and placement of them in context of presentation and differential diagnosis - Construction of a neuropsychiatric differential diagnosis - Diagnostic techniques / multi-axial classifications (including DSM based and international based systems) - Clinical or research neurophysiology (including electrodiagnostic and neuromagnetic assessment techniques) - Electrodiagnosis - Misdiagnosis in Neuropsychiatry - Diagnosis and management of neuropsychiatric aspects of movement disorders (including idiopathic Parkinson’s disease, progressive supranuclear palsy, corticobasal syndromes, dementia with Lewy Bodies, Huntington’s disease, dystonia, Tourette’s syndrome) |
| Lab tests | - Haematological, metabolic, bacteriological, virological, immunological and toxicological investigations of relevance to neuropsychiatry - Laboratory studies - Biomarkers of neurobehavioral and neuropsychiatric conditions, including neurodegeneration and neurotrauma - Indications for serum and urine studies - Indications for and interpretation of results from CSF examinations |
| Visual system assessment | - Assessment of deficits in language, praxis, gnosis, visuospatial function and other cognitive syndromes - Opthalmoscopy and perimetry |
| Forensic and risk assessment | - Forensic evaluation in patients with brain dysfunction - Risk assessment |

**Table S4.** Syllabus items for the intervention categories

| *Syllabus component* | *Syllabus item* |
| --- | --- |
| Psychological interventions | - CBT / Behaviour therapy for behavioural problems and other symptoms - Supportive therapy - Treatment, including pharmacology and other physical treatments (ECT, TMS, Surgical interventions), without neglecting psychotherapeutic and rehabilitative interventions - Family and systems therapy - Behavioural management strategies - Psychotherapies - Psychoeducation |
| Rehabilitation and neurorehabilitation | - Neuropsychiatric rehabilitation (including cognitive rehab) - Principles of neurorehabilitation and familiarity with the concepts of disability and handicap - Treatment, including pharmacology and other physical treatments (ECT, TMS, Surgical interventions), without neglecting psychotherapeutic and rehabilitative interventions |
| Pharmacological treatments | - Treatment, including pharmacology and other physical treatments (ECT, TMS, Surgical interventions), without neglecting psychotherapeutic and rehabilitative interventions - The biochemical basis of neuropsychopharmacology - Potential drug interactions between psychiatric and neurological medications and other treatments - Other medications in neuropsychiatry, including atypicals, anti-convulsants and anti-depressants - Psychotropic and neuropharmacologic agents - Neuropharmacology - Adverse effects of neuropharmacologic agents and drug-drug interactions - Neuropsychiatric aspects of psychopharmacological treatment - Psychosis and antipsychotic agents - Cognitive enhancers |
| Acute, crisis, and intensive care | - Crisis intervention in BN and NP - Consultation to intensive care |
| Somatic therapies | - Treatment, including pharmacology and other physical treatments (ECT, TMS, Surgical interventions), without neglecting psychotherapeutic and rehabilitative interventions - ECT (electroconvulsive therapy) - Understanding of newer physical treatments such as transcranial magnetic stimulation (TMS), vagus nerve stimulation (VNS), deep brain stimulation (DBS), and other physical treatments - Somatic Therapies |
| Case management and formulation | - Use of a formulation-based approach to help patients (including age, gender, culture, SES, familial factors) - Support to community teams or allied disciplines treating patients - Ongoing care for patients with functional disorders using a case management approach - Diagnosis and management of neuropsychiatric aspects of brain injury - Ability to support other professionals with challenging aspects of the neuropsychiatry of brain injury, including managing challenging behaviour and assessing mental capacity - Diagnosis and management of neuropsychiatric aspects of epilepsy - Management of the ongoing care of patients with epilepsy complicated by psychiatric comorbidity or additional functional / non-epileptic seizures - Assessment and management of treatment of patients with neuropsychiatric disorders such as those with psychiatric and behavioural symptoms and co-existing neurological disorder - Principles underlying the choice and integration of services and interventions in neuropsychiatry - Biopsychosocial management of neuropsychiatric disorders - Principles of risk assessment and management |
| Policies, practice, and frameworks | - Understanding of current guidelines and evidence base for treatments - Ability to support other professionals with challenging aspects of the neuropsychiatry of brain injury, including managing challenging behaviour and assessing mental capacity - Principles underlying the choice and integration of services and interventions in neuropsychiatry - Principles of risk assessment and management - Knowledge and appropriate application of adult safeguarding processes |
| Social and community interventions | - Support to community teams or allied disciplines treating patients - Environmental interventions - Occupational therapy - Educational therapy - Recreational therapy - Reconditioning therapy - Hydrotherapy - Prevention and health promotion in Neuropsychiatry - Biopsychosocial management of neuropsychiatric disorders |
| Single item syllabus components   - Immune therapy |  |

**Table S5.** Final data extraction sheet for syllabus coding

| *Extracted Syllabus Component* | *Present* |
| --- | --- |
| Brain injury |  |
| Epilepsy |  |
| Diagnosis and management of neuropsychiatric aspects of movement disorders (including idiopathic Parkinson’s disease, progressive supranuclear palsy, corticobasal syndromes, dementia with Lewy Bodies, Huntington’s disease, dystonia, Tourette’s syndrome) |  |
| Autoimmune disorders affecting the central nervous system, including autoimmune encephalopathies |  |
| Post-COVID syndrome |  |
| Inpatient Neuropsychiatry |  |
| General liaison neuropsychiatry |  |
| Old age liaison |  |
| (Neuro) Radiology |  |
| EEG (evoked potentials and brain-mapping) and other neurophysiological investigations and their interpretation |  |
| Complex somatic and medically unexplained symptoms |  |
| Cognitive disorders |  |
| Multiple sclerosis (and other demyelinating diseases) |  |
| Autism |  |
| Anxiety Disorders |  |
| Trauma |  |
| MDT working |  |
| Neuro-oncology |  |
| Neuromuscular disease |  |
| Understands the pathophysiology of functional symptoms |  |
| Can use a clinical assessment to make an accurate diagnosis of patients referred to a functional disorders clinic, including assessment of co-existing functional and “organic” symptoms, psychiatric co-morbidity and multi-system functional somatic symptoms |  |
| Use a formulation-based approach to help patients (including age, gender, culture, SES, familial factors) |  |
| Functional Neurological Disorder (FND) (also cited as conversion disorder) |  |
| Understanding the current guidelines and evidence base for treatments |  |
| Support to community teams or allied disciplines treating patients |  |
| Provide ongoing care for patients with functional disorders using a case management approach |  |
| Diagnosis and management of neuropsychiatric aspects of brain injury |  |
| Understands the range of cognitive and behavioural deficits associated with acquired brain injury, and the impact of these on quality of life and care-giver burden |  |
| Able to assess patients with acquired brain injury in acute inpatient and specialist outpatient settings, including neurological, cognitive and behavioural aspects of the presentation |  |
| Able to support other professionals with challenging aspects of the neuropsychiatry of brain injury, including managing challenging behaviour and assessing mental capacity |  |
| Diagnosis and management of neuropsychiatric aspects of epilepsy |  |
| Understands the range of cognitive symptoms and psychiatric co-morbidity associated with epilepsy (including mood disorders and psychosis) and anti-epileptic drugs |  |
| Able to assess patients with seizures complicated by cognitive or psychiatric symptoms and identify the contributions of seizure activity, medication and co-morbid mental health conditions to their difficulties |  |
| Able to manage the ongoing care of patients with epilepsy complicated by psychiatric comorbidity or additional functional / non-epileptic seizures |  |
| Able to use clinical skills to differentiate functional versus “organic” movement disorder, including in challenging cases where both may co-exist |  |
| Able to use clinical assessment to diagnose common psychiatric complications of movement disorders including anxiety, depression, impulse control disorders and psychosis |  |
| Acute behavioural disturbance |  |
| Acquired brain injury (both traumatic and non-traumatic) |  |
| Chronic / progressive behavioural and cognitive change |  |
| Dementia |  |
| Psychotic symptoms in patients with neurological disorders |  |
| Psychiatric disorders including depression, anxiety disorders, PTSD, emotionally unstable personality disorder |  |
| Sound knowledge base of the neuroscientific principles underlying neuropsychiatric practice (in relation to neuroanatomy, neurophysiology, neurochemistry and neuropharmacology) |  |
| Knowledge base in clinical neuroscience |  |
| Clinical skills in neuropsychiatry |  |
| Neuropsychiatric diagnosis including history and examination, neurophysiological investigations, neuroimaging, neuropsychology, and other investigations |  |
| Treatment, including pharmacology and other physical treatments (ECT, TMS, Surgical interventions), without neglecting psychotherapeutic and rehabilitative interventions |  |
| Non-dementing cognitive disorders |  |
| Seizure disorders |  |
| Traumatic brain injury (and sequelae) |  |
| Secondary psychiatric disorders, i.e. psychosis, depression, mania and anxiety disorders secondary to ‘organic’ brain disease |  |
| Substance-induced psychiatric disorders – alcohol, drugs of abuse, addiction etc |  |
| Attentional disorders (adult ADHD and related syndromes) |  |
| Developmental Neuropsychiatry |  |
| Sleep disorders |  |
| Neuropsychiatric rehabilitation (including cognitive rehab) |  |
| Forensic neuropsychiatry |  |
| Knowledge of brain structure at the macroscopic and microscopic levels, in particular the knowledge of neuronal networks, the limbic system, the neuroanatomical substrates of memory and the frontal executive system |  |
| A knowledge of CNS structure-function correlates |  |
| Knowledge of neurochemistry (especially neurotransmitter and receptor function, neurotransmitters, neuropeptides, neurohormones) |  |
| The biochemical basis of neuropsychopharmacology |  |
| The basic principles of neurophysiology |  |
| The basic principles of genetics and immunology as they apply to the CNS |  |
| A basic grasp of issues related to the mind-brain debate, the biology of consciousness and other neurophilosophical issues |  |
| Take a neuropsychiatric history; this includes all of the information routinely gathered as part of a psychiatric and medical history, screening tests and informant questionnaires |  |
| Neuropsychiatric assessment |  |
| Perform a cognitive examination (simple and extended) |  |
| Neurological examination |  |
| Assessing deficits in language, praxis, gnosis, visuospatial function and other cognitive syndromes |  |
| Interpreting neuropsychological tests |  |
| Interpret any abnormal signs elicited in neurological / medical examination and place them in context of presentation and a differential diagnosis |  |
| Construct a neuropsychiatric differential diagnosis |  |
| Understanding patterns of psychiatric symptomatology and presentation |  |
| Be familiar with the range of organic disorders |  |
| Haematological, metabolic, bacteriological, virological, immunological and toxicological investigations of relevance to neuropsychiatry |  |
| Sound understanding of the indications for, and interpretations of, the various forms of brain imaging, both structural and functional, including MRI, CT, SPECT and PET |  |
| Assess and manage treatment of patients with neuropsychiatric disorders such as those with psychiatric and behavioural symptoms and co-existing neurological disorder |  |
| Potential drug interactions between psychiatric and neurological medications and other treatments |  |
| Non-pharmacological treatments in neurological and neuropsychiatric disorders |  |
| ECT (electroconvulsive therapy) |  |
| Understanding of the newer physical treatments such as transcranial magnetic stimulation (TMS), vagus nerve stimulation (VNS), deep brain stimulation (DBS), and other physical treatments |  |
| Principles of neuro-rehabilitation and familiarity with the concepts of disability and handicap |  |
| Relationship between neuropsychiatry and allied psychiatric subspecialties such as old age, child and learning disability psychiatry |  |
| All the steps in an empirical project |  |
| Frontal / executive syndromes (of disinhibitory and non-spontaneous types) |  |
| Cognitive impairment |  |
| Diagnostic techniques / multi-axial classifications (including DSM based and international based systems) |  |
| Other medications in neuropsychiatry, including atypicals, anti-convulsants and anti-depressants |  |
| Medico-legal aspects of epilepsy |  |
| CBT / Behaviour therapy for behavioural problems and other symptoms |  |
| Structural and functional neuroanatomy (networks of psychiatric significance) |  |
| Cognition (e.g., arousal, perception, attention, language, memory, praxis recognition, visuospatial function, executive function) |  |
| Emotion (e.g., mood, affect, prosody [affective communication]) |  |
| Behavior (e.g., motivation, comportment, personality) |  |
| Mental Status Examination (and interpretation of) |  |
| Neuropsychological (psychometric) testing |  |
| Neuroimaging |  |
| Principles and applications of structural and functional imagining of the brain |  |
| Electrophysiologic Testing |  |
| Laboratory studies |  |
| Somatic Therapies |  |
| Psychotropic and neuropharmacologic agents |  |
| Neurobehavioural and Neuropsychiatric symptoms |  |
| Focal Neurobehavioural syndromes |  |
| Neuropsychiatric syndromes |  |
| Disorders of arousal (e.g., coma, vegetative states, minimally conscious states) |  |
| Perception (e.g., illusions, hallucinations, sensory impairments) |  |
| Attention (e.g., delirium / acute confusional states, hemispatial inattention / neglect) |  |
| Disorders of Language (e.g., aphasias, affective aprosodias) |  |
| (Disorders of) Memory (e.g., amnesias) |  |
| Praxis (e.g., apraxias) |  |
| Recognition (e.g., agnosias) |  |
| (Disorders of) Executive function |  |
| Social cognition (e.g., comportment, emotional recognition, theory of mind) |  |
| Aggression (and violence) |  |
| Motivation |  |
| Amnestic disorders |  |
| Disorders of mood |  |
| Disorders of affect |  |
| Factitious disorders |  |
| Malingering |  |
| Sexual disorders |  |
| Tic disorders (including Gilles de la Tourette's) |  |
| Neurodegenerative disorders |  |
| Developmental disorders |  |
| Alzheimer's disease |  |
| Frontotemporal Dementia |  |
| Lewy body disease |  |
| Parkinson's disease |  |
| Huntington's disease |  |
| Stroke (and the neuropsychiatric sequelae of) |  |
| Cerebrovascular disorders |  |
| Hydrocephalus |  |
| Primary and secondary brain tumours |  |
| CNS infections (including neuropsychiatric manifestations of HIV) |  |
| Toxic exposures / ingestions |  |
| Metabolic disorders |  |
| Headache / migraine |  |
| Acute and chronic pain (disorders) |  |
| Geriatric Behavioural Neurology and Neuropsychiatry |  |
| Pediatric BN & NP |  |
| Neuropsychiatry of substance abuse |  |
| Neurogenetics |  |
| Crisis intervention in BN and NP |  |
| Clinical or research neurophysiology (including electrodiagnostic and neuromagnetic assessment techniques) |  |
| Neuropharmacology |  |
| Neuropathology |  |
| Epidemiology, public health, public policy, and / or public advocacy |  |
| Administration / administrative psychiatry |  |
| Education |  |
| Cerebral cortex and its parts |  |
| Limbic and paralimbic structures |  |
| Basal ganglia |  |
| Brainstem(Hypothalamus, Thalamus and Internal Capsule) |  |
| Cerebellum |  |
| White matter |  |
| Cortico-cortical-subcortical circuits and networks |  |
| Regional cerebral specialization |  |
| Principles of neurology |  |
| Elemental neurological function |  |
| Neurological “soft-signs” |  |
| The use of neurological examination rating scales and the interpretation of such data |  |
| General assessment of cognition, emotion, and behavior |  |
| Standardized assessments of neuropsychiatric symptoms and syndromes |  |
| Content, sensitivity, and specificity of neuropsychological testing |  |
| Factors that influence neuropsychological test performance |  |
| Relationship between neuropsychological tests and bedside or office-based quantified clinical assessments |  |
| Correlation between neuroimaging and clinical examination |  |
| Biomarkers of neurobehavioral and neuropsychiatric conditions, including neurodegeneration and neurotrauma |  |
| Indications for serum and urine studies |  |
| Indications for and interpretation of results from CSF examinations |  |
| Adverse effects of neuropharmacologic agents and drug-drug interactions |  |
| Supportive therapy |  |
| Family and systems therapy |  |
| Environmental interventions |  |
| Behavioural management strategies |  |
| Intellectual disabilities |  |
| Communication disorders |  |
| (Specific) learning disorder |  |
| Motor disorders (including developmental coordination disorder and stereotypic movement disorder) |  |
| Obsessive-compulsive and related disorders |  |
| Trauma and stress-related disorders |  |
| Dissociative disorder |  |
| Disruptive, impulse-control and conduct disorders |  |
| Neurocognitive disorders, including delirium, mild cognitive impairment (i.e., Mild Neurocognitive Disorder) and dementia (i.e., Major Neurocognitive Disorder) |  |
| Paraphilic disorders |  |
| Personality disorders / change due to neurological conditions |  |
| Hypoxic-ischemic brain injury |  |
| (Neuro) endocrine disorders |  |
| Conducting forensic evaluations in patients with brain dysfunction |  |
| Diagnosis of delirium |  |
| Diagnosis of occupational exposure-related syndromes |  |
| Neuropsychiatric aspects of psychopharmacological treatment |  |
| Behavioural disorders (in intellectual disability, autism, and other developmental disabilities) |  |
| Catatonic behaviour and other motoric disturbances |  |
| Neurobiology |  |
| Neuropsychology of neoplastic and systemic disorders |  |
| Paediatrics neurology / neuroscience |  |
| MND |  |
| (Assessment of) Impact upon relatives and carers |  |
| Ethical codes of conduct |  |
| Medico-legal aspects to the practice of neuropsychiatry (including MHA) |  |
| Psychotherapies |  |
| Hydrotherapy |  |
| Occupational therapy |  |
| Educational therapy |  |
| Recreational therapy |  |
| Reconditioning therapy |  |
| History of neuropsychiatry |  |
| Mental hygiene |  |
| Ophthalmoscopy and perimetry |  |
| Electrodiagnosis |  |
| Encephalograms |  |
| Ventriculography |  |
| Psychobiology (biology Psychology) |  |
| Psychopathology |  |
| Psychosomatic psychiatry |  |
| Pathophysiology of neuropsychiatric disorders |  |
| Epidemiology of neuropsychiatric disorders in various populations |  |
| Phenomenology of neurologic disorders |  |
| Functional assessments |  |
| Congenital disorders |  |
| Vascular disorders |  |
| Nutritional disorders and Nutrition in Neuropsychiatry |  |
| Infections and related inflammatory disorders |  |
| Performing psychoeducation |  |
| Principles underlying the choice and integration of services and interventions in neuropsychiatry |  |
| Prevention and health promotion in Neuropsychiatry |  |
| Startle syndromes and myoclonus |  |
| Disturbances of appetite and sexual behaviour |  |
| Psychosis and antipsychotic agents |  |
| Cognitive enhancers |  |
| Psychiatric phenomenology |  |
| Psychodynamics |  |
| Conceptual, philosophical and legal issues in neuropsychiatry |  |
| Critique and synthesis of the existing scientific literature and apply this to practice |  |
| Biopsychosocial management of neuropsychiatric disorders |  |
| Statistics and research methods (including critical thinking in research and scholarship) |  |
| Neuropsychology and cognitive neuroscience |  |
| Animal Models of Neuropsychiatric diseases |  |
| Sociology of neuropsychiatry |  |
| Neurological diseases |  |
| Cognitive tests |  |
| Depersonalisation / DID |  |
| Chronic fatigue |  |
| Principles of risk assessment and management |  |
| CSF disorders |  |
| Neurosurgery |  |
| Hypnosis |  |
| Mindfulness |  |
| Misdiagnosis in Neuropsychiatry |  |
| Psychedelic therapies |  |
| Immune therapy |  |
| Knowledge and appropriate application of adult safeguarding processes |  |
| Amygdala |  |
| Disorders of Speech and Thought |  |
| Disorders of Judgment |  |
| Neurobiology of Schizophrenia |  |
| Risk assessment |  |
| Consultation to intensive care |  |
